# Pulmonary haemorrhage as a frequent cause of death among patients with severe complicated Leptospirosis in Southern Sri Lanka

**DOI:** 10.1101/2023.05.04.23289495

**Authors:** CL Fonseka, NJ Dahanayake, DJD Mihiran, AWK Mithunika, HLL Nawanjana, HS Wickramasuriya, WMDGB Wijayaratne, K Sanjaya, CK Bodinayake

## Abstract

**Background:** Leptospirosis is a tropical disease associated with life threatening complications. Identifying clinical and investigation-based parameters that predict mortality and morbidity is vital to provide optimal supportive care.

**Methods:** We conducted a prospective observational study in an endemic setting, in the southern Sri Lanka. Consecutive patients having complicated leptospirosis were recruited over 18 months. Clinical, investigational and treatment data were collected and the predictors of mortality were analysed.

**Results:** Out of 88 patients having complicated leptospirosis, 89% were male. Mean age was 47yrs (±16.0). Among the total, 94.3% had acute kidney injury, 38.6% had pulmonary haemorrhages, 12.5% had fulminant hepatic failure, 60.2% had hemodynamic instability and 33% had myocarditis. An acute significant reduction of haemoglobin (Hb) was observed in 79.4% of patients with pulmonary haemorrhage. The mean Hb reduction of patients with pulmonary haemorrhage was 3.1g/dl. The presence of pulmonary haemorrhage (PH) and hemodynamic instability within first 48 hours of admission significantly predicted mortality(p<0.05) in severe Leptospirosis. Additionally, within first 48 hours of admission, elevated SGOT, presence of atrial fibrillation, presence of significant haemoglobin reduction, higher number of inotropes used, prolonged shock, invasive ventilation and admission to ICU significantly predicted morality. Out of organ complications in the first week of illness, pulmonary haemorrhage and fulminant hepatic failure (FHF) combination had significant adjusted odds of mortality (OR=6.5 and 4.8, p<0.05). Six patients with severe respiratory failure due to PH underwent ECMO and four survived. The overall mortality in complicated leptospirosis was 17%. In PH and FHF, the mortality rate was higher reaching 35.4% and 54.5%, respectively.

**Conclusions:** Within first 48 hours of admission, organ complications such as pulmonary haemorrhage and haemodynamic instability and other parameters such as atrial fibrillation, acute haemoglobin reduction, elevated SGOT level could be used as early parameters predictive of mortality in severe Leptospirosis. PH and FHF within the first week of admission in leptospirosis are associated with high morbidity and mortality requiring prolonged ICU care and hospitalisation. Above parameters could be used as parameters indicating severity for triaging and intensifying treatment. Using ECMO is a plausible treatment option in patients with severe pulmonary haemorrhage.

**Author summary:** Leptospirosis is a tropical infectious disease predominantly affecting the lower socioeconomic groups in Sri Lanka. It is associated with significant morbidity and mortality, especially in an endemic setting. Hence, it is vital to identify clinical and biochemical parameters that can predict mortality for triaging and for escalation of care. We observed that pulmonary haemorrhage, myocarditis, hemodynamic instability and hepatic dysfunction are frequent complications of leptospirosis in southern Sri Lanka. Additionally, we identified that mortality was associated with the presence of two major complications of leptospirosis: pulmonary haemorrhage and haemodynamic instability. Therefore, early detection of these two complications along with other parameters that predict mortality such as elevated SGOT levels on admission, acute haemoglobin reduction, atrial fibrillation, prolonged shock and need for invasive ventilation would assist to recognise critically ill patients. Within the first week of admission, PH and development of FHF predicted mortality. One-third our population had acute kidney injury in isolation and they had lesser mortality. Other organ complications were almost always detected in combination and were associated with a higher mortality. Pulmonary haemorrhage was detected in one-third of patients and the majority warranted intensive care. Other than usual treatment modalities, ECMO (Extracorporeal Membrane Oxygenation) was used in six patients with critical respiratory failure due to pulmonary haemorrhage, where four survived. Out of the total group of complicated leptospirosis, in excess of one-third required intensive care treatment and 17% succumbed. Additionally, we mapped the leptospirosis prevalence rate in Galle district, and observed that severe cases are detected in specific localities. These features are helpful in early detection of severe disease and proactive management for those who are having predictors of mortality.

## Introduction

Leptospirosis has emerged as a major health threat in tropical and even in sub-tropical settings, estimated to infect more than a million people with approximately 60,000 deaths annually[1]. Natural disasters and extreme weather events including floods are now recognized to precipitate leptospirosis related epidemics [2] and its emergence in tropical settings is observed even in Sri Lanka[3]. Severe leptospirosis is characterized by dysfunction of multiple organs including the liver, kidneys, lungs, and brain. The combination of jaundice and renal failure was commonly known as Weil’s disease[4].

The major burden attributed to leptospirosis is due to its severe life-threatening manifestations such as acute renal failure, myocarditis, pulmonary haemorrhage and liver involvement. Out of the life-threatening manifestations, pulmonary haemorrhage (PH) is recognized as the most fatal. This life-threatening complication was observed to be prevalent in several leptospirosis endemic countries such as Nicaragua, El Salvador, Sri Lanka and Greece [5-8]. The case fatality for pulmonary haemorrhage syndrome was reported to be more than 50% [9], but in an endemic setting it could be as high as 100%[10-12]. A recent review noted 1.4-45.4% of lung involvement among hospitalized patients in Sri Lanka[13], and a recent study done in southern Sri Lanka mentioned an incidence just above 60%[14].

Different studies have mentioned parameters or manifestations that can be associated with mortality from Leptospirosis. Few studies have mentioned that altered mental status, oliguria, and respiratory insufficiency, hypotension, arrythmias were independently associated with death[15-17]. Out of organ related complications, acute kidney injury (AKI) is found to be the most common manifestation with a median series mortality around 10%[18]. Myocarditis was found in 7–15% of confirmed cases in Sri Lanka[19, 20]. Hepatic involvement is common and can commonly present as slight disturbances in serum liver function tests. However, fulminant hepatic failure is occasionally reported as a recognised complication in a few instances of severe leptospirosis[21]. It was evident that frequency of the complication could vary depending on the infecting serovar and geographic location.

In severe cases of pulmonary haemorrhage, some preferred using advanced therapeutic modalities such as plasmapheresis and ECMO. Although these techniques were previously reserved for developed countries, currently there are attempts to use these in developing countries where leptospirosis is endemic[22]. Currently, there is a paucity of knowledge on usage of ECMO in patients with pulmonary haemorrhage in an endemic tropical setting, only reported several case reports[22].

In our study, we describe morbidity, mortality and their predictors in a potentially severe group of leptospirosis who had organ complications, excluding relatively benign mild cases of leptospirosis without significant organ involvement. Additionally, we describe details from a large prospective case series of critically ill patients with pulmonary involvement who required ECMO.

## Methods

### Study sample

A prospective observational study was conducted at the Teaching Hospital Karapitiya (THK), Galle, the largest tertiary care centre in southern Sri Lanka, for 18 months, from March 2017 to September 2018. THK is the main referral center for complicated leptospirosis and for ECMO, due to its geographic location surrounding paddy farming communities. All consenting consecutive patients meeting the criteria for complicated leptospirosis (with at least single organ involvement) as defined below were recruited from all medical units, emergency care unit and intensive care units(ICU) of THK. The criteria included patients with documented fever or a history of fever within 7 days of admission with or without myalgia with one or more of the following: evidence of acute renal insufficiency without prerenal etiology: creatinine >1.5 mg/dL (130μmol/L) and /or urine output less than 500mL/24h with active urinary sediments (at least two of WBC, RBC and albumin on dipstick or >5 RBC/WBC per HPF), evidence of respiratory failure: Respiratory rate >28 cycles per min, evidence of hypoxia requiring oxygen, or requiring non-invasive or mechanical ventilation, evidence of spontaneous hemorrhage, pulmonary hemorrhage on chest radiograph or hemoptysis, conjunctival hemorrhage, epistaxis, hematuria, hematemesis, hematochezia or melena, unexplained vaginal bleeding, or petechiae), evidence of cardiac arrythmias or ECG changes suggestive of myocarditis or new echocardiographic abnormalities suggestive of myocarditis, evidence of hepatic dysfunction - Clinical jaundice or elevated bilirubin (total bilirubin >2mg/dL or 51.3μmol/L) or laboratory evidence of hepatitis (transaminases elevated above 2 times the upper limit of normal). Patients whose age less than 12 years, patients who are unable or unwilling to give consent for participation, leukopenia within 48 hours prior or after hospital admission, positive rapid test, virology or serology for Dengue or infections other than leptospirosis, evidence of focal bacterial infection as an aetiology of fever (i.e., urinary tract infection/ pyelonephritis, cellulitis, sinusitis, lobar pneumonia), evidence of end organ failure (renal, cardiac, respiratory) due to other etiologies unrelated to the current illness were excluded from the study.

### Definitions of complications

Oliguric renal failure was defined as urine output <0.5 ml/kg/hr and non-oliguric renal failure when urine output >0.5 mL/kg/hr. Pulmonary haemorrhage (PH) was defined from the presence of hypoxia (SpO2 <92%, paO2 < 80 mmHg/10.7 kPa) / requirement of oxygen by non-invasive or invasive methods with the presence of bilateral diffused alveolar shadows and/or drop in haemoglobin. Acute liver failure or fulminant hepatic failure was defined as the presence of PT/INR >1.5 and/or presence of hepatic encephalopathy. Cardiac involvement was defined by the presence of elevated troponin levels, changes in electrocardiogram or presence of arrythmias or echocardiographic changes. Significant acute haemoglobin reduction due to bleeding was defined as ≥2g/dl reduction of haemoglobin within 48 hours.

### Data collection

Clinical and epidemiological data were collected prospectively, using an interviewer administered questionnaire by a trained research assistant by conducting direct interviews of patients, and from hospital records after obtaining written informed consent from study participants. Laboratory and clinical parameters on first 48 hours of admission were used to analyse predictors of mortality. When the patient was in critical condition, the consent was obtained from the next-of-kin. The patients were visited every other day by the investigators to record clinical data and laboratory data. Follow-up was done by a brief assessment at the hospital performed one month after discharge from the hospital or by a telephone conversation.

### Laboratory conformation

Laboratory confirmation was based on the WHO LERG report, by symptoms consistent with leptospirosis and a single Microscopic Agglutination Test (MAT) titre ≥1:400 and/or by detection of *Leptospira* DNA by PCR and/or by the presence of IgM antibodies [23]. Leptospirosis PCR and MAT (15 pathogenic serovar panel) was performed at Medical Research Institute, Colombo, Sri Lanka.

### Spatial distribution of leptospirosis occurrence

GPS location of location of probable exposure to leptospirosis or home was recorded during the study. The spatial distribution of the patients was overlaid with the local Divisional secretariat (DS) division map of the all districts and the prevalence of leptospirosis cases per hundred thousand people in each DS division was illustrated using ArcGIS 10.3.

### Statistical testing

All data were analyzed with SPSS version 26.0. Results were expressed by mean ± standard deviation (SD) or percentages. Comparison between the two groups was performed using Pearson’s Chi-square test and Student’s T test. Mann–Whitney test was used for parameters with a non-normal distribution. A logistic regression model was used for quantitative variables for binary outcome of mortality. Unadjusted odds ratios (OR) and 95% confidence intervals (CI) were calculated. A multivariate logistic regression (stepwise forward and backward analysis) was performed to analyze the possible risk factors associated with mortality. Significance level was set at 5 % (p value ≤ 0.05).

## Results

One hundred and twenty-two patients with complicated leptospirosis were recruited to the study. Eighty-eight were eligible and the diagnosis was confirmed by PCR and/or MAT testing. Among them, mean age was 47(± 16.0) years, and 89% were male. Almost all patients had fever (97.7%) on presentation, and myalgia (81.8%), arthralgia (78.4%), vomiting (46.6%) and headache (62.5%) were other common symptoms. Eighty-three percent had a history showing exposure to leptospirosis. Thirty-two (36.4%) patients had received ICU care. Among them, the mean duration of hospitalization was 8.07(SD±5.9) days and the mean duration of ICU care was 6.73(SD±4.9) days. The case fatality rate of the study sample was 17% with 15 deaths, including one patient who succumbed on admission to the hospital.

### Complications associated with Leptospirosis (Table 1)

#### Acute kidney injury

Among the study group, acute kidney injury was the most common complication. Out of 88 patients, 83(94.3%) developed AKI. Among the group who had AKI, 73.9% had oliguric renal failure and 15.9% had non-oliguric renal failure. Out of the total, 30.7% (n=27) had only AKI.

**Table 1:**
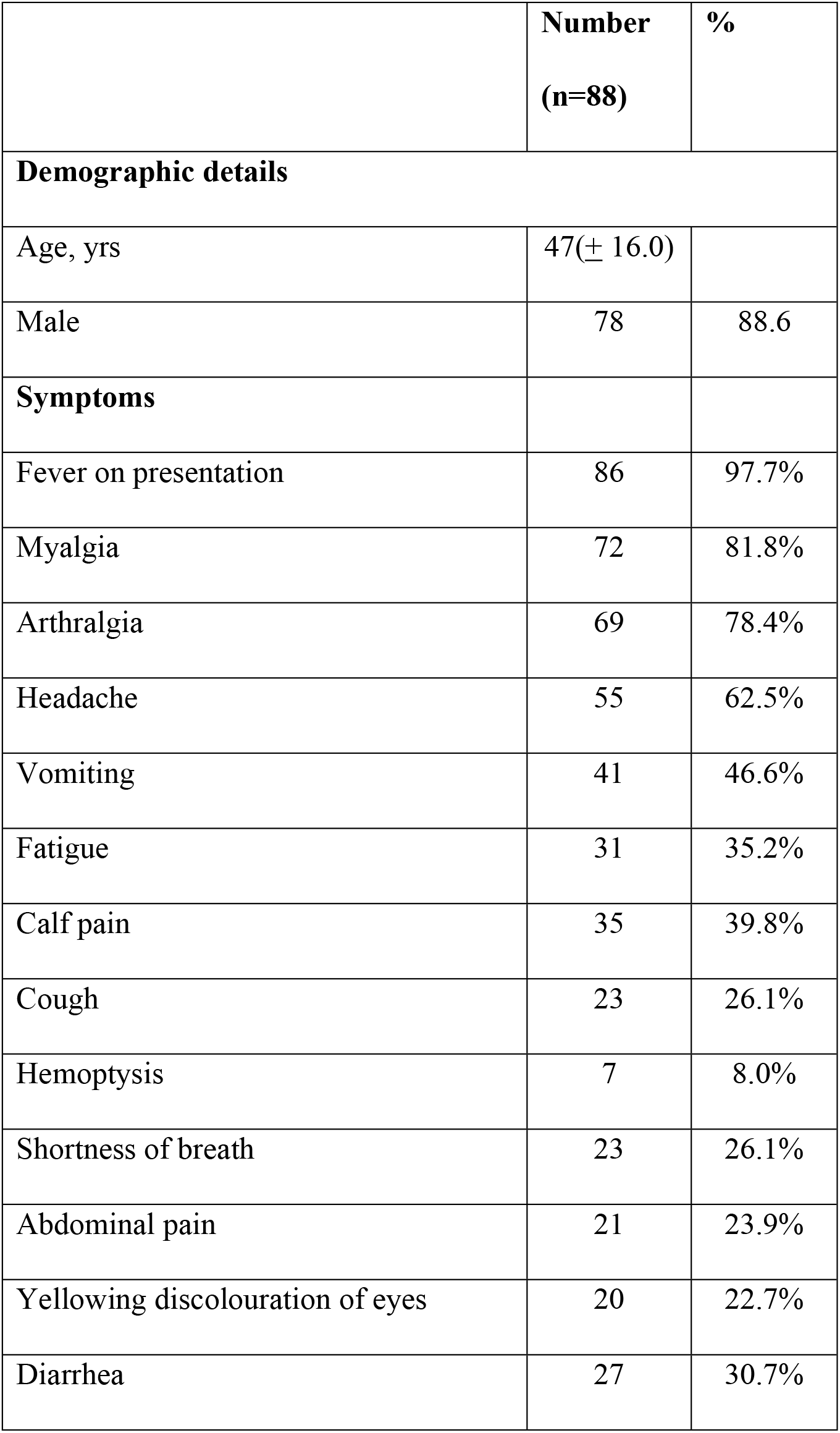
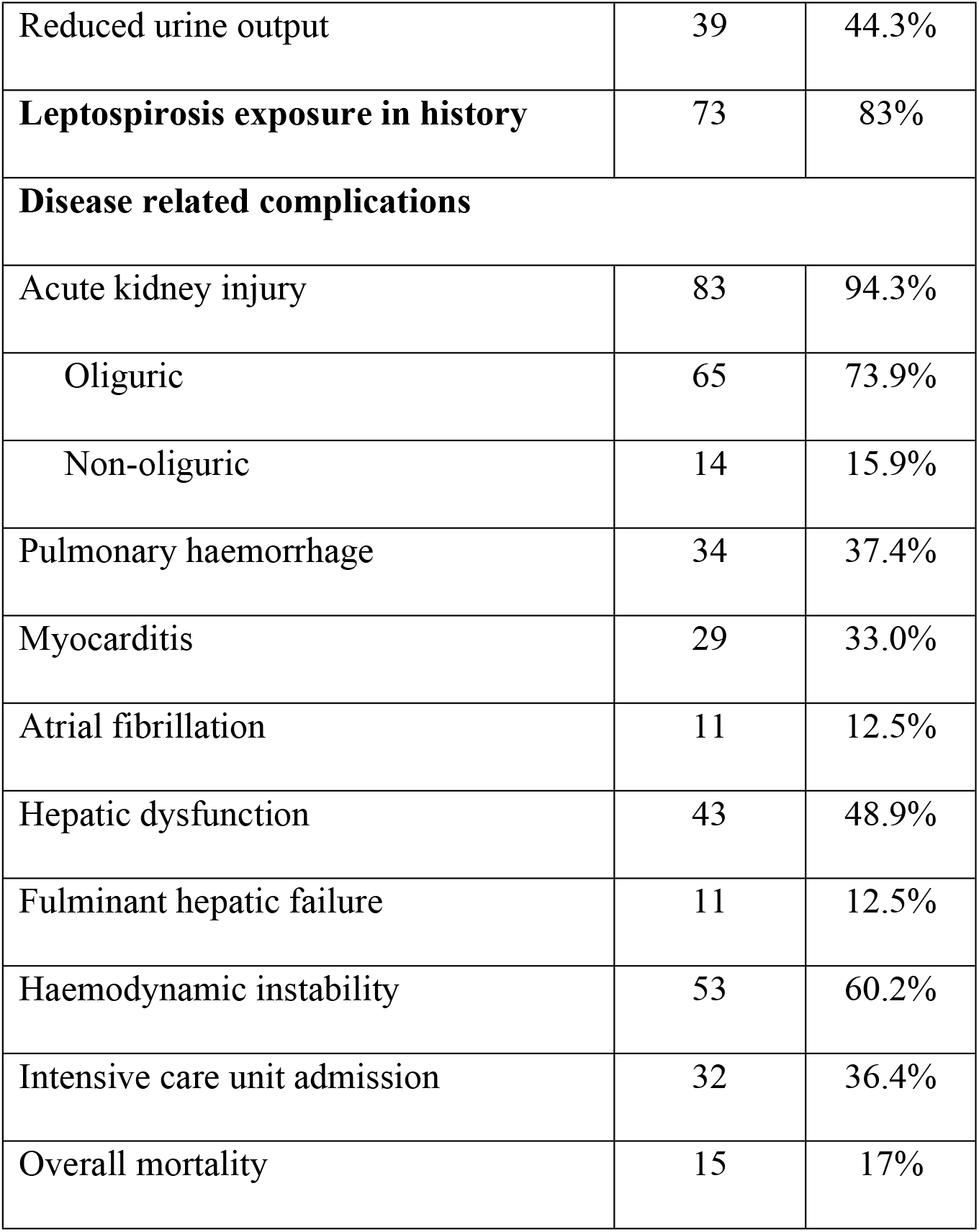
Frequency of complications in severe leptospirosis among the study group.

|  | **Number**<br>**(n=88)** | **%** |
| --- | --- | --- |
| **Demographic details** |  |  |
| Age, yrs | 47(± 16.0) |  |
| Male | 78 | 88.6 |
| **Symptoms** |  |  |
| Fever on presentation | 86 | 97.7% |
| Myalgia | 72 | 81.8% |
| Arthralgia | 69 | 78.4% |
| Headache | 55 | 62.5% |
| Vomiting | 41 | 46.6% |
| Fatigue | 31 | 35.2% |
| Calf pain | 35 | 39.8% |
| Cough | 23 | 26.1% |
| Hemoptysis | 7 | 8.0% |
| Shortness of breath | 23 | 26.1% |
| Abdominal pain | 21 | 23.9% |
| Yellowing discolouration of eyes | 20 | 22.7% |
| Diarrhea | 27 | 30.7% |
| Reduced urine output | 39 | 44.3% |
| **Leptospirosis exposure in history** | 73 | 83% |
| **Disease related complications** |  |  |
| Acute kidney injury | 83 | 94.3% |
| Oliguric | 65 | 73.9% |
| Non-oliguric | 14 | 15.9% |
| Pulmonary haemorrhage | 34 | 37.4% |
| Myocarditis | 29 | 33.0% |
| Atrial fibrillation | 11 | 12.5% |
| Hepatic dysfunction | 43 | 48.9% |
| Fulminant hepatic failure | 11 | 12.5% |
| Haemodynamic instability | 53 | 60.2% |
| Intensive care unit admission | 32 | 36.4% |
| Overall mortality | 15 | 17% |

#### Pulmonary Haemorrhage

Pulmonary haemorrhage (PH) was detected in 34(38.6%) patients. All of them had significant hypoxia and bilateral diffuse alveolar opacities. Out of them, 26.1% had cough and shortness of breath. Seven (8%) complained of haemoptysis on admission to the hospital. Twenty-four (70.6%) had bilateral diffused involvement in the chest radiograph and the rest (29.4%) had patchy bilateral alveolar shadows. Among patient who developed PH, all required supplementary oxygen, 15 (44.1%) were on non-invasive ventilation, 29 (79.4%) required intubation and ventilation.

#### Hemodynamic instability

Fifty-three (60.2%) had hemodynamic instability (hypotension with blood pressure ≤90/60) and 47 (53.4%) patients required usage of at least one inotrope during the course of treatment. Out of them, in 57.4%, 25.5%, 17% patients required one, two, three or more types of inotropes, respectively, to gain hemodynamic stability.

#### Cardiac manifestations

Out of total, 33.0% (29/88) had at least a single indicator of cardiac involvement. Elevated Troponin, abnormal ECG or an abnormality in echocardiography was detected in 18.2%, 14.8%, 11.4%, respectively. Eleven participants (12.5%) had atrial fibrillation during the illness. One had ST elevation myocardial infarction detected on admission.

#### Hepatic dysfunction

Out of total, 43 (48.9%) patients had hepatic dysfunction evidenced by deranged transaminases, ALP, total bilirubin and/or PT/INR. Thirty (34.1%) had elevated ALT and/or AST and 8 had elevated alkaline phosphate levels at the first 48 hours of the admission. Deranged PT/INR was observed at a later stage of the illness. Eleven (12.5%) patients had evidence of fulminant hepatic failure late (after 48 hours of the illness) during the course of illness. All of these individuals who fulfilled the definition of acute hepatic failure had prolonged hypotension and were supported on multiple inotropes.

#### Haematological parameters – acute haemoglobin reduction and platelet count

A significant reduction in haemoglobin (Hb) of >2g/dl within 48 hours was observed in 27 out of 34 (79.4%) having PH with a mean maximum Hb reduction of 3.1g/dl (±1.2, range = 1.3-5.9g/dl) over 48 hours. Median day of detection of a significant haemoglobin drop was on day 5 after onset of fever (Q1=4, Q3=6 days). In the non-pulmonary haemorrhage group, a minority (16.7%) had significant haemoglobin reduction. The mean significant haemoglobin reduction in this group was 1.15g/dl (±1.09, range= 0-4.5). Median day of detection of a significant haemoglobin drop in the non-PH group was on day 6 after onset of fever (Q1=4.75, Q3=8.75 days). The maximum mean haemoglobin reduction was significantly higher in the PH group than non-PH group (p=<0.001). Furthermore, significant hemoglobin reduction in non-PH group has occurred later than the PH group (p=0.046).

Among the whole group, 44 (50.0%) and 70 (79.5%) had platelet counts less than 50,000cm^3^ and less than 100,000 cm^3^, respectively.

#### Association of the number of major complications with mortality in severe Leptospirosis

Then, we analysed the association of major complications in patients with severe Leptospirosis within the first week after admission. Acute kidney injury, myocarditis, pulmonary hemorrhage, hepatic dysfunction and hemodynamic instability were considered as major complications.

**Table 2** reveals the association of major complications among confirmed leptospirosis patients. Twenty-nine (33.0%) patients had at least one complication associated with leptospirosis. Out of them, nearly all (27 patients, 93.1%) had isolated acute kidney injury, one had isolated pulmonary haemorrhages and another had isolated hemodynamic instability mimicking septic shock. In the group of patients who had only two major complications, majority (n=13, 68.4%) had acute kidney injury with hemodynamic instability. Increasing number of complications significantly increased the admission to ICU, duration of hospital stays and mortality (p=<0.05).

**Table 2:**
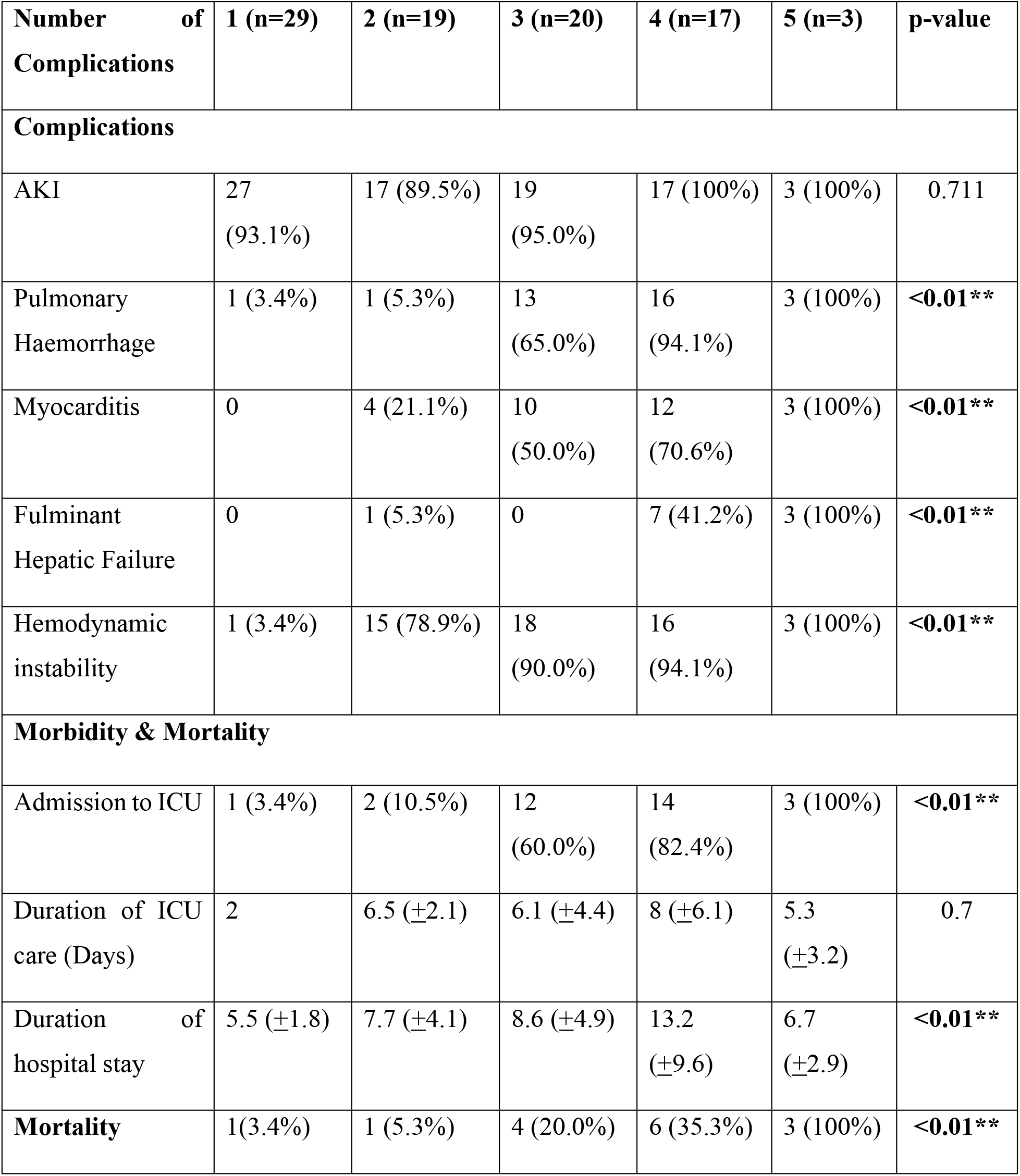
Morbidity and Mortality in patients having major complications of severe Leptospirosis.

| Number of Complications | 1 (n=29) | 2 (n=19) | 3 (n=20) | 4 (n=17) | 5 (n=3) | p-value |
| --- | --- | --- | --- | --- | --- | --- |
| **Complications** |  |  |  |  |  |  |
| AKI | 27 (93.1%) | 17 (89.5%) | 19 (95.0%) | 17 (100%) | 3 (100%) | 0.711 |
| Pulmonary Haemorrhage | 1 (3.4%) | 1 (5.3%) | 13 (65.0%) | 16 (94.1%) | 3 (100%) | **<0.01\*\*** |
| Myocarditis | 0 | 4 (21.1%) | 10 (50.0%) | 12 (70.6%) | 3 (100%) | **<0.01\*\*** |
| Fulminant Hepatic Failure | 0 | 1 (5.3%) | 0 | 7 (41.2%) | 3 (100%) | **<0.01\*\*** |
| Hemodynamic instability | 1 (3.4%) | 15 (78.9%) | 18 (90.0%) | 16 (94.1%) | 3 (100%) | **<0.01\*\*** |
| **Morbidity & Mortality** |  |  |  |  |  |  |
| Admission to ICU | 1 (3.4%) | 2 (10.5%) | 12 (60.0%) | 14 (82.4%) | 3 (100%) | **<0.01\*\*** |
| Duration of ICU care (Days) | 2 | 6.5 (±2.1) | 6.1 (±4.4) | 8 (±6.1) | 5.3 (±3.2) | 0.7 |
| Duration of hospital stay | 5.5 (±1.8) | 7.7 (±4.1) | 8.6 (±4.9) | 13.2 (±9.6) | 6.7 (±2.9) | **<0.01\*\*** |
| **Mortality** | 1(3.4%) | 1 (5.3%) | 4 (20.0%) | 6 (35.3%) | 3 (100%) | **<0.01\*\*** |

#### Early predictors of mortality in patients with severe Leptospirosis

Clinical and biochemical parameters within first 48 hours of admission were considered for analysis. Symptoms present on admission were not predictive of mortality, which included symptoms such as haemoptysis, shortness of breath, abdominal pain etc. However, the presence of atrial fibrillation, presence of a significant acute haemoglobin reduction, increase number of inotropes used, prolonged shock, intubation and ventilation, and admission to ICU significantly predicated morality in the severe leptospirosis group. Although, platelet, white cell count, serum bilirubin during admission were not predictive of mortality, high SGOT level on admission (but not SGPT) level significantly predicted mortality (Table 3).

**Table 3:**
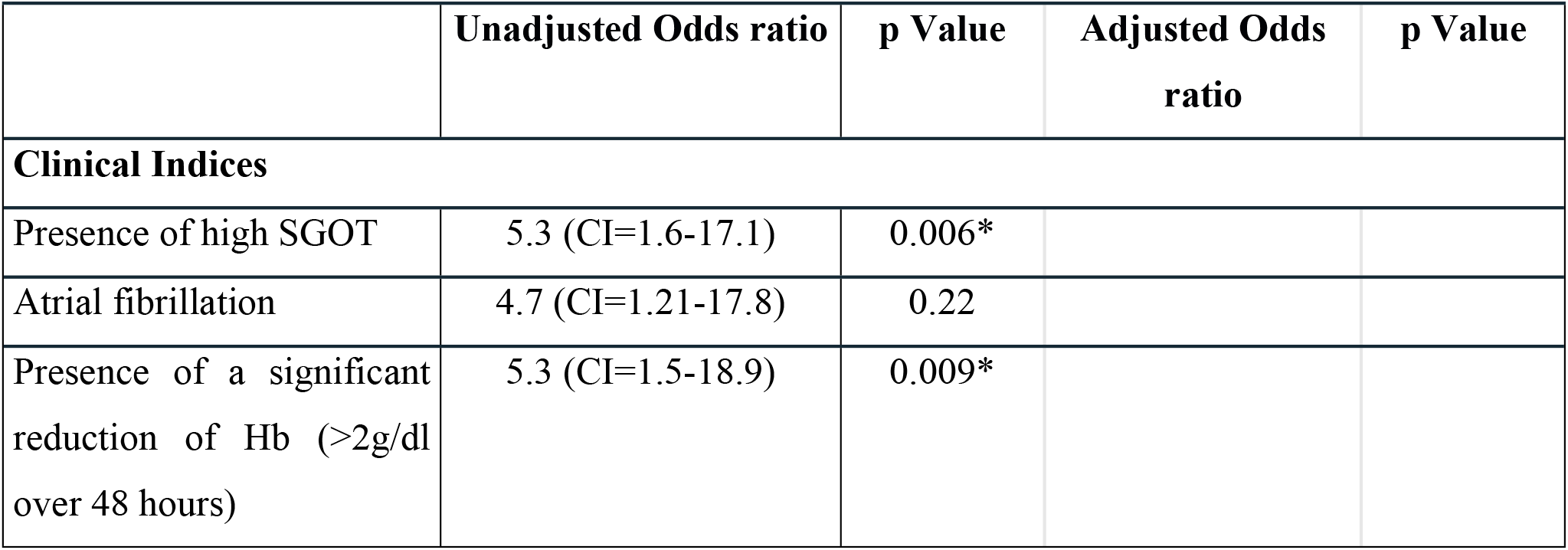
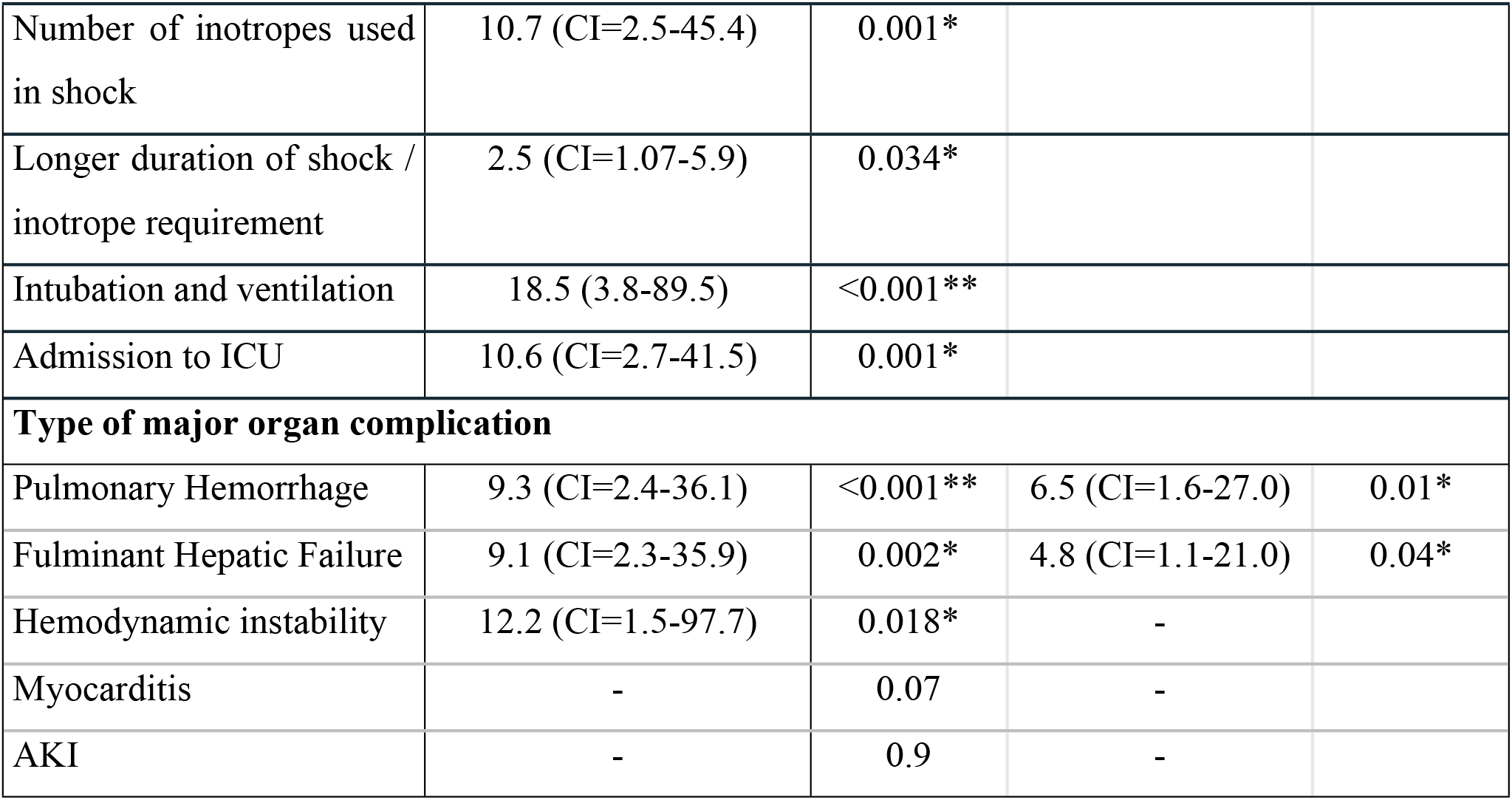
Clinical indices and Major organ complications predictive of mortality in patients with severe leptospirosis. (Logistic regression analysis of predictors of mortality)

|  | Unadjusted Odds ratio | p Value | Adjusted Odds ratio | p Value |
| --- | --- | --- | --- | --- |
| **Clinical Indices** |  |  |  |  |
| Presence of high SGOT | 5.3 (CI=1.6-17.1) | 0.006* |  |  |
| Atrial fibrillation | 4.7 (CI=1.21-17.8) | 0.22 |  |  |
| Presence of a significant reduction of Hb (>2g/dl over 48 hours) | 5.3 (CI=1.5-18.9) | 0.009* |  |  |
| Number of inotropes used in shock | 10.7 (CI=2.5-45.4) | 0.001* |  |  |
| Longer duration of shock / inotrope requirement | 2.5 (CI=1.07-5.9) | 0.034* |  |  |
| Intubation and ventilation | 18.5 (3.8-89.5) | <0.001** |  |  |
| Admission to ICU | 10.6 (CI=2.7-41.5) | 0.001* |  |  |
| **Type of major organ complication** |  |  |  |  |
| Pulmonary Hemorrhage | 9.3 (CI=2.4-36.1) | <0.001** | 6.5 (CI=1.6-27.0) | 0.01* |
| Fulminant Hepatic Failure | 9.1 (CI=2.3-35.9) | 0.002* | 4.8 (CI=1.1-21.0) | 0.04* |
| Hemodynamic instability | 12.2 (CI=1.5-97.7) | 0.018* | - |  |
| Myocarditis | - | 0.07 | - |  |
| AKI | - | 0.9 | - |  |

#### Association of organ involvement with mortality in severe Leptospirosis

We analysed whether these five major complications were predictive of mortality in patients with severe Leptospirosis (Table 3). Due to the long hospital stay in certain individuals the complications within the first week of stay in hospital was considered for this analysis. The presence of pulmonary haemorrhage (PH), fulminant hepatic failure (FHF) and hemodynamic instability significantly predicts mortality(p=<0.05). Additional analysis with stepwise logistic regression showed that, out of the five complications, PH and FHF combination were most predictive of mortality.

#### Comparison of mortality and morbidity in patients having Pulmonary Haemorrhage (PH)

**Table 4** depicts a comparison of clinical characteristics and morbidity and mortality among patients who had pulmonary haemorrhage compared with those who did not.

**Table 4:**
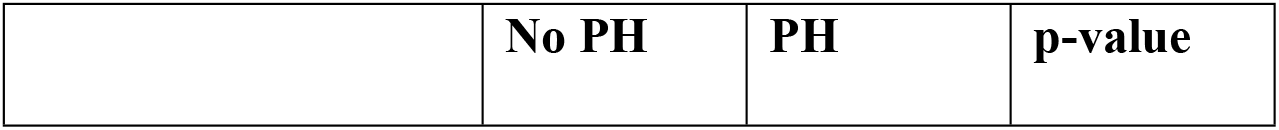
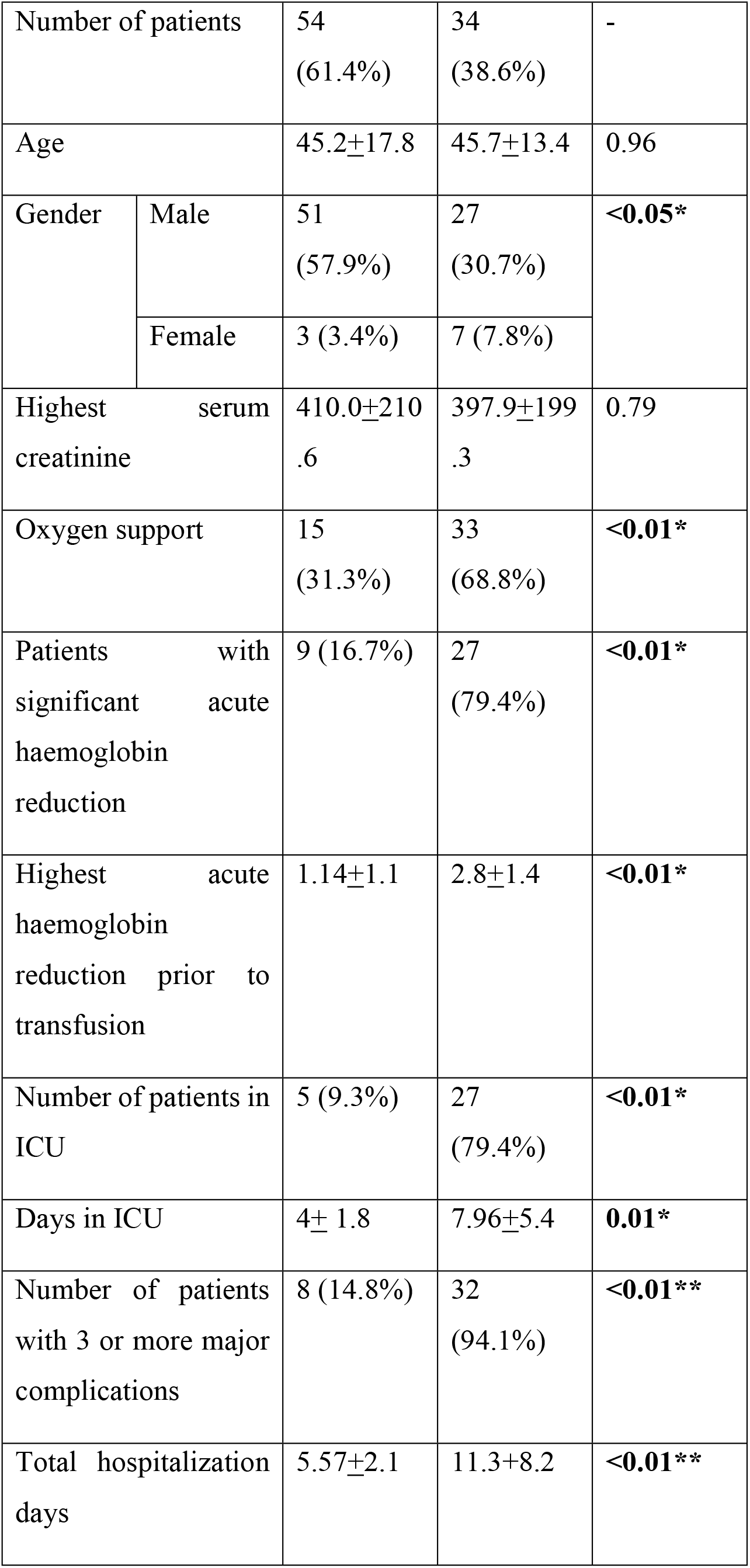
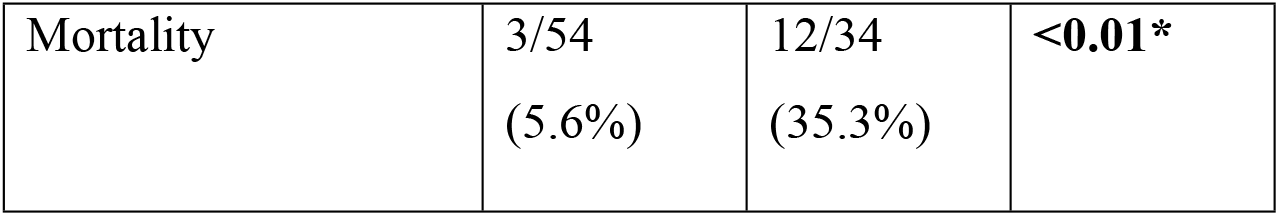
Characteristics of complications and morbidity among patients having pulmonary haemorrhage (PH) in severe Leptospirosis.

|  |  |  |  |  |
| --- | --- | --- | --- | --- |
| Number of patients |  | 54<br>(61.4%) | 34<br>(38.6%) | - |
| Age |  | 45.2±17.8 | 45.7±13.4 | 0.96 |
| Gender | Male | 51<br>(57.9%) | 27<br>(30.7%) | <0.05* |
|  | Female | 3 (3.4%) | 7 (7.8%) |  |
| Highest serum creatinine |  | 410.0±210.6 | 397.9±199.3 | 0.79 |
| Oxygen support |  | 15<br>(31.3%) | 33<br>(68.8%) | <0.01* |
| Patients with significant acute haemoglobin reduction |  | 9 (16.7%) | 27<br>(79.4%) | <0.01* |
| Highest acute haemoglobin reduction prior to transfusion |  | 1.14±1.1 | 2.8±1.4 | <0.01* |
| Number of patients in ICU |  | 5 (9.3%) | 27<br>(79.4%) | <0.01* |
| Days in ICU |  | 4± 1.8 | 7.96±5.4 | 0.01* |
| Number of patients with 3 or more major complications |  | 8 (14.8%) | 32<br>(94.1%) | <0.01** |
| Total hospitalization days |  | 5.57±2.1 | 11.3±8.2 | <0.01** |

|  |  |  |  |
| --- | --- | --- | --- |
| Mortality | 3/54<br>(5.6%) | 12/34<br>(35.3%) | **<0.01\*** |

This shows that females had a significantly higher risk of developing PH than males. The PH group required significantly higher non-invasive ventilation, intubation and ICU care and had a higher number of days in ICU and in hospitalization. Out of the patients who had PH, all most all had two or more major complications and had a significantly higher mortality rate compared to the group who had no PH (35.3% vs 5.6%). Nearly 80% patients with PH required management in an ICU. Among the haematological parameters, PH group (71,400/μl ±46,600) had significantly lower mean platelets counts on admission (104,000μl ±69,300) (p=0.03). However, white blood cell count on admission was not significantly between the two groups.

#### Non-invasive and invasive treatment modalities used in patients with severe Leptospirosis

Next, we analysed the treatment modalities used in patients with severe Leptospirosis. All patients received antibiotics. Penicillin, ceftriaxone and doxycycline were the most frequently used antibiotics either alone or in combination: Penicillin or Ceftriaxone monotherapy for 27.3%, penicillin and doxycycline for 13.6%, ceftriaxone and doxycycline for 38.6%. Methylprednisolone was administered to 56(63.6%) and intravenous immunoglobulin (IV Ig) was given to 22(25.0%) patients. Tranexamic acid was given to 49(55.7%) patients.

Sixteen (18.2%) patients required dialysis. Two patients required continuous renal replacement therapy (CRRT) concurrently during ECMO. Supplementary oxygen to 55.7%, NIV (CPAP) to 21.6% and intubation and ventilation to 36.4%, were required. Plasmapheresis was performed in 39(44.3%) and 6 (6.6%) required ECMO.

#### ECMO in patients with Leptospirosis

ECMO was required in 6 (6.6%) patients with severe pulmonary haemorrhage. Eligibility of ECMO was decided on the severity of respiratory failure calculated by the ‘Murray score’ which consist of PaO2/FiO2 (P/F) ratio, extent of alveolar involvement on chest radiograph and peak end-expiratory pressure (PEEP) in the ventilator setting[24], physician’s discretion and the presence of other co-morbidities. All patients had evidence of severe respiratory failure with Murray scores of 3.25 or above and poor oxygenation despite high ventilator settings. They had a mean P/F (paO2/FiO2) ratio of 93 before ECMO. All had dense bilateral diffused opacities in the chest radiograph (Supplementary: Figure 1a,b). All underwent veno-venous ECMO and were supported by ECMO for a mean duration of 151.4 hours (± 55.83). They were intubated and ventilated for a mean of 12 days (±5.70) and were in ICU for a mean 14.8 ±5.44 days (total of 118 days). All required renal replacement therapy (RRT). Serial chest radiographs of these patients typically showed resolution of alveolar shadows within 4 to 5 days after initiation of ECMO (Supplementary: Figure 1c,d,e). Four got completely cured and were well at 1- and 3-month follow-up. Two patients succumbed despite ECMO therapy (Supplementary: Table 1).

#### Overall morbidity and mortality in patients with Leptospirosis

Overall, the mean duration of hospitalization was 8.07(SD±5.9) days. Thirty-two (36.4%) intubated and ventilated and received ICU care for a mean of 6.73 (±4.9) days. Out of them, 17 received ICU care for <7days, 4 for 7-14days and 6 for more than 2 weeks. The patients were intubated and ventilated for mean days of 5.97 (SD±4.8) days.

The case fatality rate of the study sample was 17% with 15 deaths. Patients who developed PH and FHF had a mortality rate of 35.2% and 54.5%, respectively.

The fatality rate of patient with pulmonary haemorrhage who has undergone plasmapheresis is 32.3%. Direct comparison for efficacy was not made as the majority (91.2%) of patients with pulmonary haemorrhage have undergone plasmapheresis and only 3 in PH group did not receive plasmapheresis.

#### Spatial distribution of the abundance of Leptospirosis cases

Total leptospirosis cases, Leptospirosis cases with pulmonary haemorrhage and the number of deaths due to Leptospirosis were mapped for Galle district of Sri Lanka. Figure 1 illustrates that Elpitiya and Yakkalamulla DS divisions are the hot spots for overall leptospirosis cases and Leptospirosis cases with pulmonary haemorrhage (Figure 1 (a) and (b)). On the contrary, Neluwa DS division had the highest death rate due to leptospirosis among all divisions (Figure 1 (c)).

**Figure 1:**
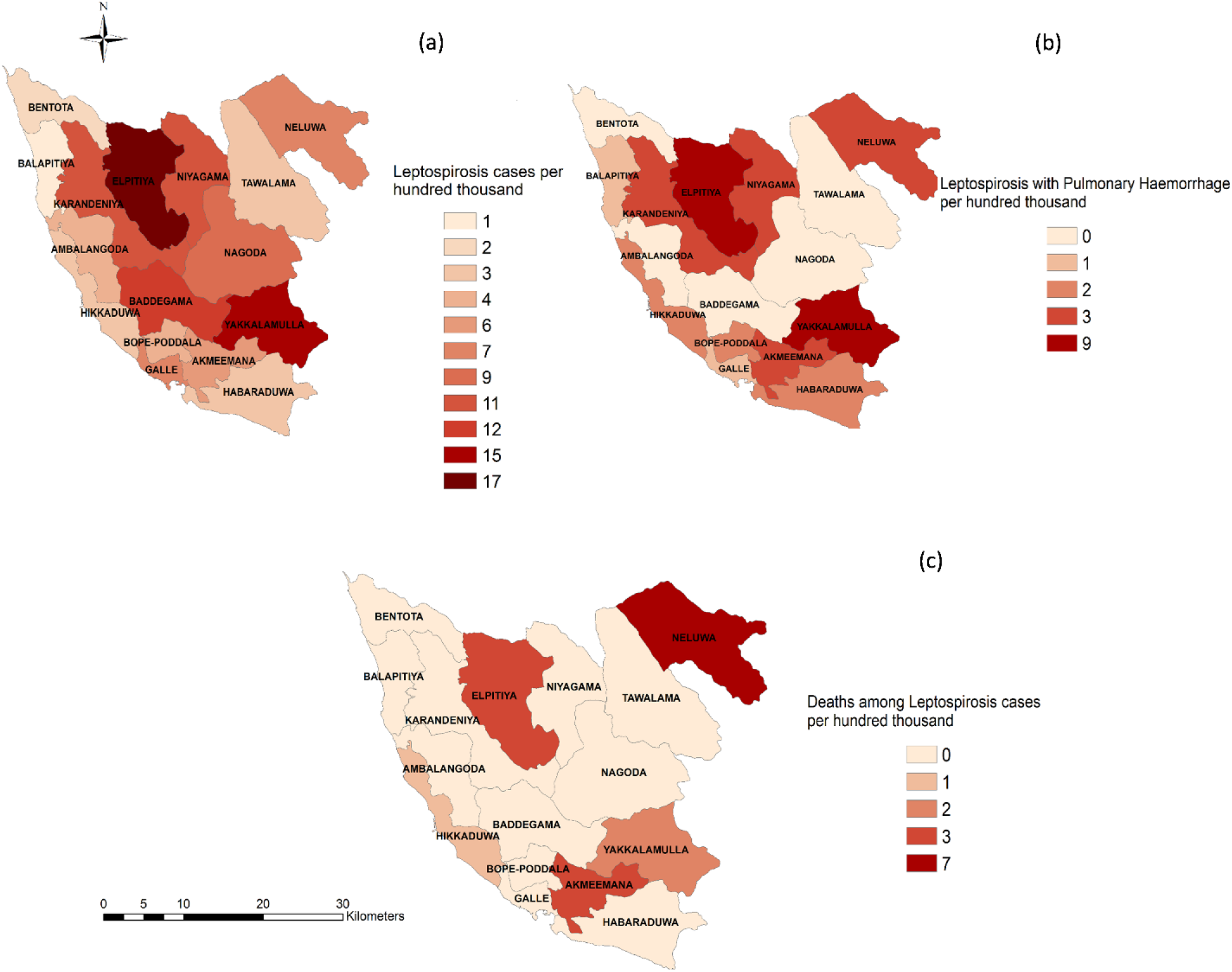
The spatial distribution of the cases and deaths of leptospirosis in Galle district, Sri Lanka.

## Discussion

In this prospective study, we observed that pulmonary hemorrhage (PH) and hemodynamic instability early in the presentation were significantly associated with mortality. In Galle, PH were present in one-third and FHF was present in 12.5% of patients with severe leptospirosis. We observed that 80% of patients requiring ICU admission had pulmonary hemorrhage, warning us of the significant morbidity in this group. Hepatic involvement with FHF is considered as an unusual clinical manifestation of severe leptospirosis[25, 26], and has only been reported in several instances[21]. Features of fulminant hepatitis were not present early in the illness and developed in patient who require inotropic support which indicate that the pathogenesis could be related to ischaemic liver injury than direct hepatic involvement. However, the presence indicated risk of morbidity and mortality. Therefore, other than the routinely performed hematological tests and renal function tests, we suggest that focused examination and investigations should be performed for early detection of PH and FHF. We suggest to perform prothrombin time (INR), transaminases, hemoglobin reduction, chest radiograph, monitor for hypoxia to identify these complications early. AKI presented as the only manifestation in the majority and other organ complications usually associate with AKI. The presence of a single organ related complication is associated with lesser mortality and with each addition of each organ related complication an increase in mortality is observed. Additionally, we have identified hot spots area of leptospirosis cases and deaths are prevalent in Galle district. This would give us more understanding on how to triage these patients that require strict monitoring and supportive care.

Septic shock due to leptospirosis is an under-recognised complication. We also noticed that hemodynamic instability is observed in two-thirds of patients with severe leptospirosis. Most of them required inotropic support, where 50% required 2 or more inotropes. It was observed that the septic shock in Leptospirosis is much difficult to manage than septic shock due to other conventional bacterial organism. However, comparative studies on clinical and cytokine correlated will be of value to understand the pathogenesis. It has been suggested that delay in treating patients with antibiotics and intravenous hydration was an important factor in mortality[28]. Fluid management in severe leptospirosis is more challenging than bacterial sepsis due to the co-occurrence of AKI, myocarditis or multiple organ dysfunction syndrome. Additionally, we should be aware that myocardial dysfunction and severe bleeding can mimic septic shock. Therefore, it has been suggested that a more conservative fluid management approach may be more appropriate in complicated leptospirosis than managing according to standard sepsis guidelines, given the high mortality associated with pulmonary involvement etc[29]. Yilmaz et al. examined ICU patients and found that the clinical and laboratory findings of leptospirosis are similar to those of sepsis. They recommended to think about leptospirosis while examining a patient with SIRS/sepsis etiology in an area endemic for leptospirosis[30]. In a study done in a tropical Australian setting, they have suggested that prompt ICU support, early antibiotics, conservative fluid resuscitation, protective ventilation strategies, traditional thresholds for RRT initiation, and corticosteroid therapy, associated with a very low case-fatality rate[31]. Given the high incidence of hemodynamic instability in our group, we should be more vigilant that septic shock, bleeding and myocarditis should be identified promptly and managed accordingly.

Although, we could not find any symptoms present on admission or hematological parameters other than SGOT, that can predict mortality; atrial fibrillation, significant acute hemoglobin reduction, longer duration of shock, requirement of intubation and ICU care were important clinical indicators predicting mortality. It has been reported that disproportionate exaggerated rise of SGOT associated with grave prognosis in the late phase of leptospirosis in a limited number of patients[32]. However, we observed that SGOT can predict mortality at an early phase up to 5 days after the onset of fever.

There is a dearth of evidence regarding the extent of hemoglobin reduction detectable in patients with leptospirosis associated pulmonary hemorrhage. We observed that in the PH group, the majority had a significant hemoglobin reduction on 4-6 days after symptom onset. We also observed a mean maximum hemoglobin reduction of 5.6g/dl in patients with PH. This hemoglobin reduction was observed despite efforts to immediately resuscitate and transfuse patients having acute bleeding. Due to the fact that acute hemoglobin reduction predicted mortality, we suggest that it is advisable to cross match 4-5 units of blood upon suspicion of PH. Moreover, in a leptospirosis endemic area of Brazil, they have noted that 13% had gastrointestinal bleeding while a similar percentage developed pulmonary hemorrhages[27]. Therefore, it is plausible that other than pulmonary hemorrhages, leptospirosis could lead to concealed bleeding elsewhere, judging from the fact that significant hemoglobin reduction was observed in nearly 15% of patients who did not have features to suggest pulmonary hemorrhages in our study.

As we have only included patients with one or more complication related to leptospirosis, we consider that these patients represent a potentially severe group that needs hospital care and advanced treatment modalities. Moreover, our study includes a large prospective case series of leptospirosis who underwent ECMO in the world. Altogether, there were 11 reported individual cases and two retrospective studies mentioning 13 patients with ARDS due to leptospirosis[22, 32-34] who underwent ECMO. Patients in our study were selected for ECMO when the Murray score was 3-4. ECMO improved oxygenation in these patients who had inadequate oxygenation despite being on maximum ventilatory support. We described 6 patients who underwent ECMO and four survived. Respiratory failure occurring due to pulmonary hemorrhages are transient and reversible, whereas the chances of survival may be improved if they are supported adequately during this critical period. Initiation of ECMO enabled to maintain adequate oxygenation because the lungs could be maintained on ‘rest settings’ at low ventilatory pressures until haemorrhagic lungs recover. Therefore, ‘rest settings’ potentially could reduce inotrope requirement due to restoration of venous return and tissue perfusion due to lesser ventilatory pressures. Due to prevalent septic shock clinical phenotype requiring multiple inotropes, theoretically ECMO would assist to stabilise haemodynamic parameters initially. Traditionally, severe bleeding has been a relative contraindication to extra-corporeal membrane oxygenation (ECMO), which requires systemic anticoagulation to maintain circuit patency. However, cases of diffuse alveolar haemorrhage have been reported to be successfully managed using ECMO[35]. In our case series, standards levels of ACT were used and any aggravation of bleeding was not observed. This should be used cautiously in patients with PH and anticoagulation should be withheld immediately with exacerbation of bleeding. In a tropical setting where funding and resources are sparse, we consider this a major step forward to improve mortality in patients with severe pulmonary haemorrhage due to Leptospirosis.

Ideally, further observational studies would have been beneficial to decide on surrogate respiratory parameters (Murray score, P/F ratio, oxygenation index) where ECMO can be initiated. This would invite the need for further research or trials to observe a benefit of ECMO and plasma exchange in patients with pulmonary haemorrhage due to Leptospirosis.

## Conclusions

Pulmonary haemorrhage and haemodynamic instability can be used as early predictors of mortality in severe leptospirosis. Other than the above atrial fibrillation, acute haemoglobin reduction, prolonged shock, elevated SGOT level at admission can be used as warning signs of mortality. These findings will have a significant effect on the management of patients in taking decisions on triaging and escalation of care. Identifying disease hot spots of leptospirosis cases and deaths will help in decisions to improve regional hospital facilities and organised transfer for advanced care.

## Data Availability

Raw data will be available in Faculty of Medicine, University of Ruhuna website in a data repository.

## Ethical approval

Ethical approval for the study was obtained from the Ethical Review Committee, Faculty of Medicine, University of Ruhuna. Approval for conducting the research at THK was obtained from the Director of the THK. Informed consent was taken from all study participants.

## Author contributions

CLF, NJD, CKB are joint senior authors involved in conceptualisation, conducting and editing of the manuscript. SK analysed the GPS data and created figures. Other authors contributed in patient recruitment, data collection, data analysis and writing initial draft. All authors contributed significantly to this work.

## Acknowledgement

We would like to thank the ECMO team: Dr T Harischandra, Dr K Withanaarachchi and Dr RK Firmin who did an incredible service to facilitate ECMO support and Dr T Weliwita who performed plasma exchange. We are grateful to Dr Lilani Karunanayake and the Medical Research Institute for performing confirmatory investigations to diagnose leptospirosis and also sharing the investigation data with us.

